# A descriptive analysis of 2020 California Occupational Safety and Health Administration COVID-19-related complaints

**DOI:** 10.1101/2021.12.06.21262384

**Authors:** Marilyn D Thomas, Ellicott C. Matthay, Kate A. Duchowny, Alicia R. Riley, Harmon Khela, Yea-Hung Chen, Kirsten Bibbins-Domingo, M. Maria Glymour

## Abstract

COVID-19 mortality disproportionately affected specific occupations and industries. The Occupational Safety and Health Administration (OSHA) protects the health and safety of workers by setting and enforcing standards for working conditions. Workers may file OSHA complaints about unsafe conditions. Complaints may indicate poor workplace safety during the pandemic. We evaluated COVID-19-related complaints filed with California (Cal)/OSHA between January 1, 2020 and December 14, 2020 across seven industries. To assess whether workers in occupations with high COVID-19-related mortality were also most likely to file Cal/OSHA complaints, we compared industry-specific per-capita COVID-19 confirmed deaths from the California Department of Public Health with COVID-19-related complaints. Although 7,820 COVID-19-related complaints were deemed valid by Cal/OSHA, only 627 onsite inspections occurred and 32 citations were issued. Agricultural workers had the highest per-capita COVID-19 death rates (402 per 100,000 workers) but were least represented among workplace complaints (44 per 100,000 workers). Health Care workers had the highest complaint rates (81 per 100,000 workers) but the second lowest COVID-19 death rate (81 per 100,000 workers). Industries with the highest inspection rates also had high COVID-19 mortality. Our findings suggest complaints are not proportional to COVID-19 risk. Instead, higher complaint rates may reflect worker groups with greater empowerment, resources, or capacity to advocate for better protections. This capacity to advocate for safe workplaces may account for relatively low mortality rates in potentially high-risk occupations. Future research should examine factors determining worker complaints and complaint systems to promote participation of those with the greatest need of protection.

## Introduction

COVID-19 mortality disproportionately affected specific occupations and industries (Chen et al., 2021). One year into the pandemic, California had the highest total number of confirmed COVID-19 cases and deaths of any US state (3.67 million and 59,000, respectively) (The New York Times, 2021). Within the state, overall mortality during the pandemic increased 39% among food and agricultural workers, compared with 19% among health and emergency workers, and 11% among non-essential workers (Chen et al., 2021). This excess mortality indicates the risks encountered by workers in essential jobs and the potential failure to enact and enforce workplace safety measures during the pandemic. Variation in excess mortality across occupations underscores the critical role that employers play in safeguarding their workers, especially those employed in “essential sectors” (State of California, 2020a, 2020b).

Many factors might have contributed to the breakdown of workplace safety during the pandemic, but the role of the worker complaint response system merits special scrutiny. Previous studies indicate that examining occupational health and safety complaint and citation records within the workplace setting may serve as a unique data source in helping to identify risky workplace settings that have inadequate worker protections (Hanage et al., 2020; Mendeloff & Gray, 2005; Mendeloff & Seabury, 2013; Weil & Pyles, 2005). Health and safety complaints may serve as a structural indicator of exposure to SARS-CoV-2, the virus that causes COVID-19, due to the lack of worker protections. A recent time-series study using national Occupational Safety and Health Administration (OSHA) complaint data found that complaint counts predicted COVID-19 cases 10 days later and deaths 16 days later (Hanage et al., 2020). This study suggested that OSHA complaints may be used as an “early warning system” for emerging COVID-19 hotspots. OSHA complaints would be useful to guide preventive actions only if workers are able and willing to assess and report COVID-19-related occupational hazards.

OSHA protects the health and safety of working people by setting and enforcing standards for working conditions (Michaels & Wagner, 2020). Prior to guidelines imposed in 2021 (Cal/OSHA, 2021a), California employers were mandated to protect workers from COVID-19 under two pre-pandemic guidelines – the Aerosol Transmissible Diseases Standard and the Injury and Illness Prevention Program (Cal/OSHA, 2021c). Preexisting policies also required employers to report all COVID-19-related workplace fatalities or injuries like any other occupational event (Cal/OSHA), although temporary changes in face coverings and physical distancing regulations were made in June 2020 (Cal/OSHA, 2021a). To this end, employees are able to file a COVID-19-related complaint against any employer alleged to be non-compliant with the California Division of Occupational Safety and Health (Cal/OSHA) (Cal/OSHA, 2021b). Any employer who fails to implement and enforce mandated guidelines and/or correct past workplace hazards are in violation of the state and could be issued a citation by Cal/OSHA for non-compliance.

During the pandemic, thousands of COVID-19-related worker complaints were submitted to Cal/OSHA across all industries. Though dozens of citations were issued to employers deemed to be non-compliant upon investigation by the state (CA Department of Industrial Relations), determinants of complaint and inspection frequency is not straightforward. For instance, complaint frequency likely reflects some combination of true workplace hazards/risk and worker comfort/power to report risks. As such, fewer complaints in certain industries may indicate a lack of worker engagement with the Cal/OSHA system rather than a lack of workplace risk. Similarly, the frequency of onsite inspections may be influenced by agency resources that vary by timing or region as well as the severity of worker risk. An examination of the Cal/OSHA complaint system may help identify non-compliant industries and employers at greater risk for COVID-19 exposure or mortality.

In this manuscript, we describe the distribution of COVID-19-related Cal/OSHA complaints across industries during 2020 by evaluating complaint rates and characteristics (e.g., severity). To assess whether workers who are at greatest COVID-19 mortality risk are also those who report the most workplace risks, we compare industry-specific COVID-19 confirmed deaths with COVID-19-related complaints.

## Materials and Methods

We obtained data from Cal/OSHA’s Division of Occupational Safety and Health on all complaints regarding establishments allegedly violating COVID-19 safety guidelines filed between January 1, 2020 and December 14, 2020. Each COVID-19-related complaint contained information associated with the establishment (e.g., name, address, industry code), the alleged violation (e.g., complaint date, severity), employer compliance (e.g., satisfactory correction), and Cal/OSHA’s response (e.g., onsite inspection, citation issued). A complaint is affirmed by Cal/OSHA as valid if the alleged violation is determined to have a reasonable basis in fact. Complaints could be marked as invalid for myriad reasons including a pending correction to the problem or it was filed in the wrong jurisdiction. Nonformal complaints (reported by an anonymous employee) are investigated by phone or letter (CA Department of Industrial Relations). Formal complaints (reported by or on behalf of a named employee) require an onsite inspection (CA Department of Industrial Relations). Upon investigation, establishments deemed non-compliant can be issued an order of corrective action and/or a citation (i.e., a monetary fine) (CA Department of Industrial Relations). Unsatisfactory employer responses can lead to increased fines over time.

Using 2017 data from the North American Industry Classification System (NAICS) (US Census Bureau), we matched each establishment’s 6-digit industry code to its corresponding 2-digit code to obtain macro-level industrial categories. To calculate rates, we identified the number of workers associated with each industry code by summing the person weights for adults aged 18 to 65 in the 2019 California American Community Survey by 2-digit NAICS code. Last, using publicly available Cal/OSHA data on establishments receiving a citation (CA Department of Industrial Relations), we matched each citation’s inspection number to its respective proposed penalty. All data sources are publicly available hence our study was exempt from IRB approval by the Human Research Protection Program at the University of California San Francisco.

We restricted our analysis to valid Cal/OSHA complaints. We assessed all available complaint characteristics: business type (local government, private, state government); complaint type (health, safety, health & safety); severity (low, high); formality (formal, nonformal); inspection conducted (no, yes); and reason for inspection (required, follow-up, Area Director’s discretion). Statistical differences between complaints and industries were assessed using chi-square tests (p<.05). Complaint rates were calculated as the number of COVID-19-related complaints filed within a given industry per 100,000 workers. Additional statistical analyses of inspection-related factors were restricted to complaints receiving an onsite inspection. Methods for coding all complaint variables are presented in **Appendix A**.

Potential temporal alignment between COVID-19-related complaints and outcomes was evaluated using California death certificates. Individual death records were obtained from the California Department of Public Health. Each death certificate contained information on the cause of death, date of death, and decedent occupation, among other data. We assigned NAICS industry codes to the entries in the open text field for decedent occupation using an automated system developed by the National Institute for Occupational Safety and Health (NIOSH Industry and Occupation Computerized Coding System (NIOCCS)). Confirmed 2020 COVID-19 mortality deaths among those aged 18-65 were identified using the ICD-10 classification code for cause of death (U07). Like complaint rates, death rates were calculated as the number of COVID-19-confirmed deaths within a given industry per 100,000 workers.

## Results

Of the 8,736 COVID-19-related complaints made through December 14, 2020, 7,820 complaints were deemed valid by Cal/OSHA (CA Department of Industrial Relations). Five of twenty industry categories comprised two-thirds of all complaints. Therefore, we assessed 5 industries individually: Health Care & Social Assistance, Retail Trade, Manufacturing, Accommodation & Food Services, and Transportation & Warehousing. Despite comprising only 2% of complaints, we also evaluated the Agriculture, Forestry, Fishing & Hunting (hereafter Agricultural) industry given mounting evidence of high rates of infection among California agricultural workers (Chen et al., 2021; Waltenburg et al., 2021). The remaining 14 industries (range 0.1–4.6% of complaints) were collapsed into one category for analysis (‘Other’). Thus, we evaluated a total of 7 industry categories.

**Table 1** shows the distribution of COVID-19-related deaths (n=23,233) and valid Cal/OSHA complaints (n=7,820) in 2020 by industry. The total number of industry workers was 18,106,047. The overall complaint rate was 57 per 100,000 (100K) workers with the highest number of complaints among private organizations (92%). Overall complaints were largely health-related (89%) followed by health- and safety-related (10%), of lower severity (82%), nonformal (91%), and elicited no onsite inspection (92%). Though all formal complaints (reported by a named employee) typically require an inspection, only 245 of 713 (34%) received one during 2020. Among the total number of 627 inspections conducted, 382 (61%) were from nonformal complaints (reported by an anonymous employee) (**Appendix B**). On average, the death rate attributed to COVID-19 was 115 per 100K workers.

**Table 1.** Distribution of COVID-19-related California deaths and Cal/OSHA complaint characteristics by industry, 2020.

|  | Total | Health Care<br>& Social<br>Assistance | Retail<br>Trade | Manufacturing | Accommodation<br>& Food Services | Transportation<br>&<br>Warehousing | Agriculture,<br>Forestry,<br>Fishing &<br>Hunting | Other |
| --- | --- | --- | --- | --- | --- | --- | --- | --- |
|  | *row n (%)* | *row n (%)* |  |  |  |  |  |  |
| **Deaths** |  |  |  |  |  |  |  |  |
| Total | 23,233 (100) | 1,812 (7.8) | 1,532 (6.6) | 2,791 (12.0) | 1,021 (4.4) | 1,231 (5.3) | 1,458 (6.3) | 13,388 (57.6) |
| **Complaints** |  |  |  |  |  |  |  |  |
| Total | 7,820 (100.0) | 1,815 (23.2) | 1,246 (15.9) | 861 (11.0) | 723 (9.3) | 562 (7.2) | 149 (1.9) | 2,464 (31.5) |
|  | *column n (%)* | *column n (%)* |  |  |  |  |  |  |
| **Business Type** |  |  |  |  |  |  |  |  |
| Local | 438 (5.6) | 111 (6.1) | 2 (0.2) | 3 (0.3) | 1 (0.1) | 54 (9.6) | 0 (0.0) | 267 (10.8) |
| Private | 7,228 (92.4) | 1,682 (92.7) | 1,244 (99.8) | 857 (99.5) | 722 (99.9) | 506 (90.0) | 149 (100.0) | 2,068 (83.9) |
| State | 154 (2.0) | 22 (1.2) | 0 (0.0) | 1 (0.1) | 0 (0.0) | 2 (0.4) | 0 (0.0) | 129 (5.2) |
| **Complaint Type** |  |  |  |  |  |  |  |  |
| Health (H) | 6,922 (88.5) | 1,636 (90.1) | 1,124 (90.2) | 718 (83.4) | 637 (88.1) | 507 (90.2) | 143 (96.0) | 2,157 (87.5) |
| Safety (S) | 146 (1.9) | 26 (1.4) | 18 (1.4) | 15 (1.7) | 6 (0.8) | 17 (3.0) | 2 (1.3) | 62 (2.5) |
| H & S | 752 (9.6) | 153 (8.4) | 104 (8.3) | 128 (14.9) | 80 (11.1) | 38 (6.8) | 4 (2.7) | 245 (10.0) |
| **Severity** |  |  |  |  |  |  |  |  |
| Low | 6,439 (82.3) | 1,259 (69.4) | 1,106 (88.8) | 763 (88.6) | 611 (84.5) | 492 (87.5) | 121 (81.2) | 2,087 (84.7) |
| High | 1,381 (17.7) | 556 (30.6) | 140 (11.2) | 98 (11.4) | 112 (15.5) | 70 (12.5) | 28 (18.8) | 377 (15.3) |
| **Formality** |  |  |  |  |  |  |  |  |
| Formal | 713 (9.1) | 213 (11.7) | 93 (7.5) | 63 (7.3) | 55 (7.6) | 59 (10.5) | 23 (15.4) | 207 (8.4) |
| Nonformal | 7,107 (90.9) | 1,602 (88.3) | 1,153 (92.5) | 798 (92.7) | 668 (92.4) | 503 (89.5) | 126 (84.6) | 2,257 (91.6) |
| **Inspection** |  |  |  |  |  |  |  |  |
| No | 7,193 (92.0) | 1,662 (91.6) | 1,148 (92.1) | 780 (90.6) | 660 (91.3) | 521 (92.7) | 117 (78.5) | 2,305 (93.5) |
| Yes | 627 (8.0) | 153 (8.4) | 98 (7.9) | 81 (9.4) | 63 (8.7) | 41 (7.3) | 32 (21.5) | 159 (6.5) |
| **Rates** |  |  |  |  |  |  |  |  |
| *per 100,000 workers* |  | *per 100,000 workers* |  |  |  |  |  |  |
| Death Rate | 115.4 | 80.8 | 83.2 | 174.1 | 73.9 | 133.7 | 401.5 | 146.8 |
| Complaint Rate | 56.9 | 80.9 | 67.6 | 53.7 | 52.3 | 61.0 | 43.8 | 25.8 |

*Note: Chi-square tests comparing each complaint characteristic across the six industry categories were significant ( $p < .001$ ). Rates were calculated using the number of workers in a given industry: Total=18,106,047; Health Care & Social Assistance=2,243,345; Retail Trade=1,842,446; Manufacturing=1,602,690; Accommodation & Food Service=1,381,409; Transportation & Warehousing=920,798; Agriculture, Forestry, Fishing & Hunting=363,181; Other=9,752,178. Descriptions of variables and coding are detailed in Supplemental Table S1.*

The number of workers in a given industry varied widely: Health Care & Social Assistance=2,243,345; Retail Trade=1,842,446; Manufacturing=1,602,690; Accommodation & Food Service=1,381,409; Transportation & Warehousing=920,798; Agricultural=363,181; and Other=9,752,178. Complaints from workers in the health care sector accounted for the largest proportion of all complaints (23%) and highest complaint rate (81 per 100K). The health care sector had the second lowest death rate (81 per 100K). Health Care complaints were also the most likely to be rated as severe (31%) followed by Agricultural worker complaints (19%) (e.g., crop and animal production). Agricultural complaints were the most likely to be formal (15%) and exclusively health-related (96%). Death rates were highest for Agricultural workers (402 per 100K) although the sector had amongst the lowest rate of worker complaints (44 per 100K). The total number of inspections exceeded the total number of formal complaints filed in industries with highest death rates: Agricultural and Manufacturing. Conversely, formal complaints were fewer than inspections in Health Care that had the lowest death rate.

Fifteen employers had the highest number of alleged COVID-19-related violations (≥30) (**Table 2**). Three in five of these establishments were in the Health Care and Retail industries. Among the three establishments that were cited, fines ranged from $560 to $78,300. For every 10 complaints filed, Kaiser Permanente (an integrated managed health care organization) received nearly 3 on-site inspections followed by about 2 inspections for both Amazon and Ralphs, a Southern California based supermarket chain owned by Kroger Co.

**Table 2.** Characteristics of 15 establishments with the highest number of Cal/OSHA COVID-19-related complaints, 2020.

| Establishments | Industry | Number of Complaints | Inspections per 10 Complaints | Citations per 10 Inspections | Lowest Fine | Highest Fine |
| --- | --- | --- | --- | --- | --- | --- |
| Kaiser Permanente | Health Care & Social Assistance | 186 | 3 | 1 | \$1,535 | \$78,300 |
| UPS | Transportation & Warehousing | 81 | 0 | — |  |  |
| McDonalds | Accommodation & Food Services | 62 | 1 | 0 |  |  |
| Walmart | Retail Trade | 62 | 0 | — |  |  |
| Sutter Health | Health Care & Social Assistance | 59 | 2 | 0 |  |  |
| Amazon | Transportation & Warehousing | 54 | 2 | 2 | \$560 | \$935 |
| CA Corrections and Rehabilitation | Public Administration | 53 | 1 | 0 |  |  |
| Target | Retail Trade | 53 | 1 | 0 |  |  |
| FedEx | Transportation & Warehousing | 47 | 0 | — |  |  |
| CA DMV | Public Administration | 41 | 1 | 0 |  |  |
| Lowe's Home Improvement | Retail Trade | 40 | 1 | 0 |  |  |
| Alameda Health System | Health Care & Social Assistance | 37 | 0 | — |  |  |
| Dignity Health | Health Care & Social Assistance | 35 | 1 | 0 |  |  |
| Ralphs | Retail Trade | 32 | 2 | 1 | \$13,500 | \$13,500 |
| Home Depot | Retail Trade | 30 | 1 | 0 |  |  |

*Note: All complaints are investigated by Cal/OSHA but not all investigations result in an onsite inspection. All establishments that receive a citation are issued a monetary fine (proposed penalty).*

Among the employers that received an onsite inspection (n=627), 32 citations were issued (**Appendix B**). Apart from the complaint type and formality, there were significant differences in inspection characteristics by industry. Health Care had the highest proportion of inspections (24%), many from severe complaints (41%), with almost 1 in 10 resulting in a fine. Though lower in severity (16%), Agriculture employers were issued nearly the same proportion of fines (9%). Conversely, Accommodation & Food Service inspections were reportedly low in severity (89%) and no citations were issued for the 63 inspections.

Among the 32 citations issued, fines ranged from $560 to $105,000 with some establishments being cited multiple times (**Table 3**). Nearly half of fines were issued within the Health Care sector (47%) followed by the Manufacturing sector (19%). Total fines were the highest for Kaiser Permanente ($258,470) followed by Overhill Farms Incorporated ($191,850). Only one citation was issued without an onsite inspection ($55,850 to Smithfield Foods Inc).

**Table 3.** Characteristics of the 32 establishments issued a citation by Cal/OSHA for COVID-19-related complaints, 2020.

| Establishment | Industry | Reason for Inspection | Fine Issued |
| --- | --- | --- | --- |
| Overhill Farms, Inc. | Manufacturing | AD Discretion | \$105,000 |
| Overhill Farms, Inc. | Manufacturing | Required | \$86,850 |
| Kaiser Permanente | Health Care & Social Assistance | AD Discretion | \$78,300 |
| Hollywood Presbyterian Medical | Health Care & Social Assistance | Required | \$57,120 |
| Kaiser Permanente | Health Care & Social Assistance | Required | \$56,000 |
| Smithfield Foods Inc. | Wholesale Trade | No inspection | \$55,850 |
| Kaiser Permanente | Health Care & Social Assistance | Required | \$55,350 |
| Central valley Meat Holding Co. | Manufacturing | Required | \$50,000 |
| Kaiser Permanente | Health Care & Social Assistance | AD Discretion | \$39,685 |
| DL Poultry, Inc. | Manufacturing | AD Discretion | \$36,000 |
| San Miguel Villa | Health Care & Social Assistance | AD Discretion | \$32,000 |
| Primex Farms, LLC | Agriculture, Forestry, Fishing & Hunting | AD Discretion | \$27,500 |
| NorthBay Medical Center | Health Care & Social Assistance | AD Discretion | \$25,250 |
| Harvest Farms | Agriculture, Forestry, Fishing & Hunting | AD Discretion | \$23,000 |
| Marin General Hospital | Health Care & Social Assistance | AD Discretion | \$21,000 |
| Pitman Family Farms | Manufacturing | Required | \$18,000 |
| Kaiser Permanente | Health Care & Social Assistance | AD Discretion | \$16,400 |
| The Ridge Post-Acute | Health Care & Social Assistance | Required | \$15,400 |
| Food 4 Less | Retail Trade | Required | \$15,000 |
| Dragonfruit Holdings | Health Care & Social Assistance | Required | \$13,500 |
| Ralphs | Retail Trade | AD Discretion | \$13,500 |
| Kaiser Permanente | Health Care & Social Assistance | Required | \$11,200 |
| Olson Meat Company | Manufacturing | AD Discretion | \$9,000 |
| Santa Clarita Transit | Public Administration | AD Discretion | \$6,750 |
| Supermercado Mi Tierra | Retail Trade | Required | \$6,750 |
| Uni-Kool Partners | Agriculture, Forestry, Fishing & Hunting | Required | \$5,850 |
| Santa Clara Valley Medical Center | Health Care & Social Assistance | AD Discretion | \$2,060 |
| Kaiser Permanente | Health Care & Social Assistance | Required | \$1,535 |
| Amazon | Transportation & Warehousing | Required | \$935 |
| DHL | Transportation & Warehousing | Required | \$750 |
| St. Rose Hospital | Health Care & Social Assistance | Required | \$750 |
| Amazon | Transportation & Warehousing | AD Discretion | \$560 |

Note: AD = Area Director.

As shown in **Figure 1**, weekly COVID-19-related complaints spiked in mid-March 2020 and again in July, roughly preceding the first and second waves of the pandemic, respectively. Lag time in deaths was estimated by counting the number of weeks between peaks in complaints and deaths during each wave. An overall rise in deaths lagged behind the first spike by 6 weeks and behind the second spike by 3 weeks (4.5 weeks on average). The sharp third surge in deaths beginning in mid-November was not preceded by increases in occupational complaints, however.

**Figure 1.**
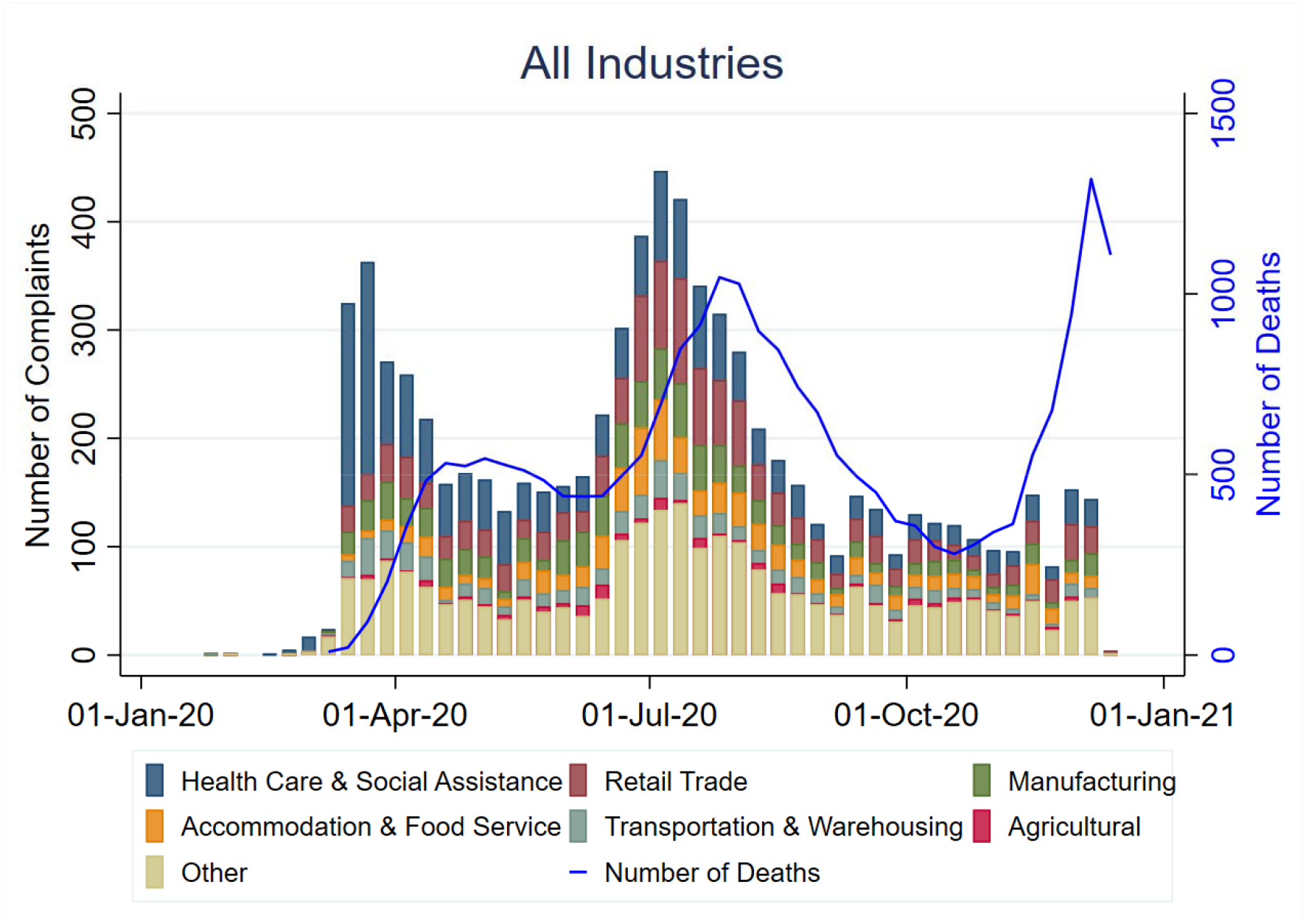
The distribution of weekly Cal/OSHA COVID-19-related complaints and COVID-19-confirmed deaths across 7 macro-level industries. Each week of 2020 is indicated along the x-axis. The multicolored stacked bar graph shows the number of COVID-19-related complaints filed within a given industry each week, as indicated along the left y-axis in black. The blue line graph represents the number of COVID-19 confirmed deaths across all industries each week, as indicated along the right y-axis in blue.

The number of complaints in Health Care far exceeded all other industries during the first wave of infection, and then lowered steadily through the second wave. Retail, Manufacturing, and Accommodation & Food Service complaints were low during the first wave and increases during wave two. Similarly, ‘other’ industries showed the largest combined increase during wave two.

Agricultural and Transportation & Warehousing complaints remained relatively low across all waves.

**Figure 2 (a-f)** illustrates weekly deaths and complaints within various occupational sectors of six industries. During the first wave, the vast majority of weekly complaints were filed by hospital workers, which greatly reduced by mid-April. By the second wave, the number of complaints were highest among machinery, store retailers, food service, and ambulatory workers. Weekly complaints were fewer than 15 among Agricultural workers and filed mostly by employees that support the industry (e.g., farm labor contractors, soil preparation, planting, crop harvesting). The number of weekly deaths were similarly patterned across most industries except Accommodation & Food and Transportation & Warehousing which trended lower. However, Agricultural worker complaints and deaths were fairly normally distributed (i.e., spikes between waves were barely discernable) until the third wave of infection in November. On average, a rise in deaths lagged behind each spike by nearly 4 weeks.

**Figure 2.**
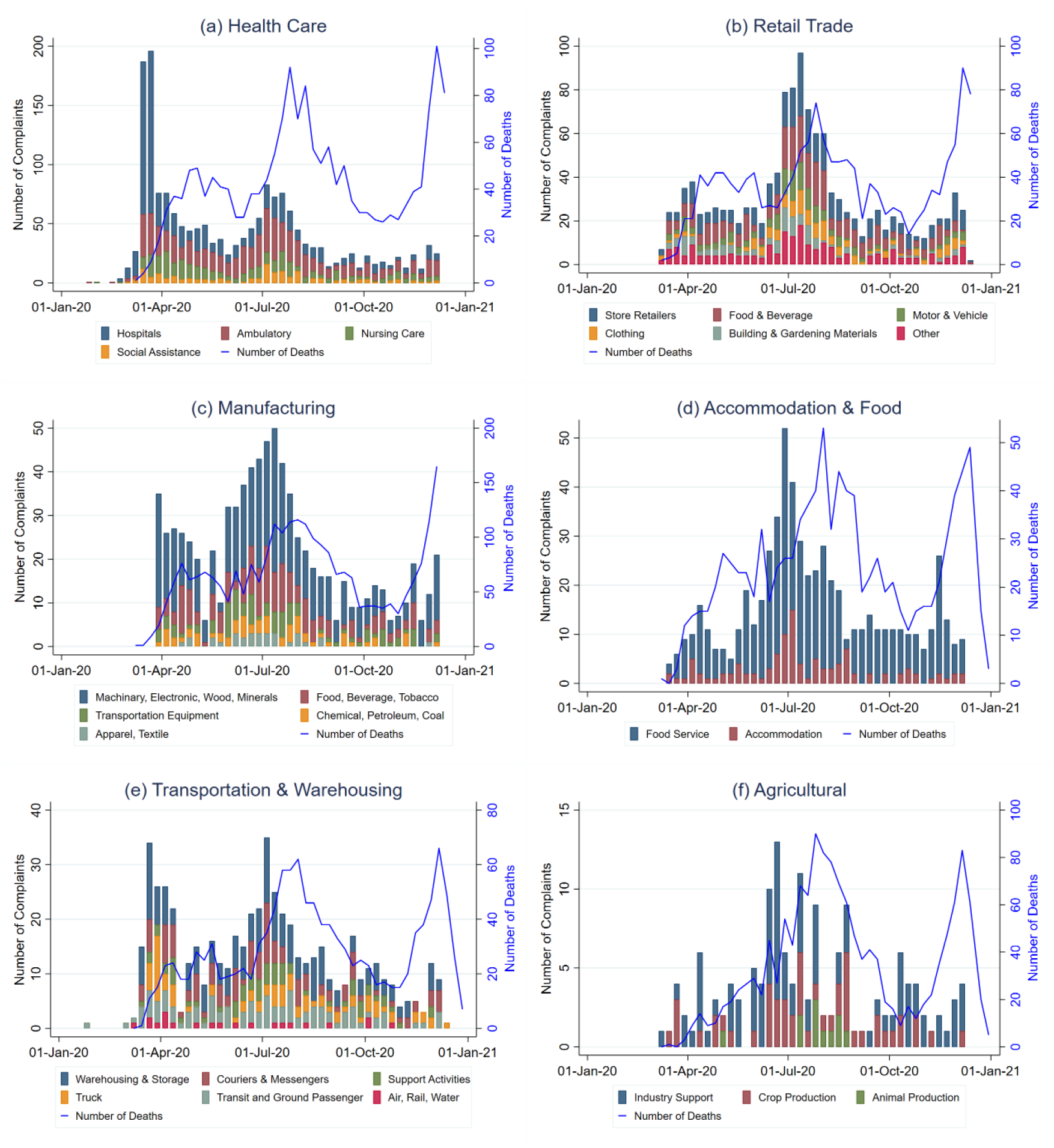
The distribution of weekly Cal/OSHA COVID-19-related complaints and COVID-19-confirmed deaths within various occupational sectors of 6 macro-level industries; (a) Health Care, (b) Retail, (c) Manufacturing, (d) Accommodation and Food Service, (e) Transportation and Warehousing, and (f) Agricultural industries. Each week of 2020 is indicated along the x-axis. The multicolored stacked bar graph shows the number of COVID-19-related complaints filed within a given industry sector each week, as indicated along the left y-axis in black. The blue line graph represents the number of COVID-19 confirmed deaths within the industry each week, as indicated along the right y-axis in blue.

## Discussion

Using all valid COVID-19 occupational safety complaints filed in the state of California during the first year of the pandemic, we found that complaints were common but rarely triggered inspections or citations. The industries with the highest COVID-19 mortality rates were not the industries from which complaints were most likely to originate. Although spikes in complaint counts preceded the first two major mortality surges, occupational complaints did not provide any early warning for the third wave of the pandemic starting in late fall.

To our knowledge, this is the first study documenting the correspondence between industry-specific OSHA complaints and industry-specific mortality rates. The variations in complaint rates across industry highlight specific sectors that may lack worker resources or protections, require greater enforcement, or need additional technical assistance to support compliance. We found that Agricultural workers had the highest COVID-19 death rates but were least represented among complaints of workplace risk, whereas Health Care workers had the highest complaint rates, the most citations and the highest fines, but had the second lowest COVID-19 death rates. These discrepancies raise questions about why industries with some of the highest COVID-19 death rates have some of the lowest complaint levels. Although high rates of COVID-19 mortality in a given worker group are not necessarily *due to* workplace exposures or violations, our findings suggest that complaints may not adequately capture COVID-19 exposure or mortality risk. The absence of complaints from worker groups that are known to be at high-risk of COVID-19 exposure, infection, and mortality, such as agricultural and manufacturing workers, is concerning, and warrants further investigation to determine if violations have gone unreported.

Another reason the Cal/OSHA complaint system may not have prevented excess mortality is inadequate enforcement, namely insufficient levels of inspections or response to worker complaints. The number of OSHA inspectors has steadily decreased in recent years, contributing to fewer overall and planned (i.e., non-complaint related) inspections thereby weakening worker protections (Brown, 2015; Mendeloff & Seabury, 2013). For example, Cal/OSHA’s inspector-to-worker ratio was nearly half that of federal OSHA in 2014.(Brown, 2015) Prior to the pandemic, Cal/OSHA conducted about 7,300 total workplace safety inspections annually among private employers (OSHA). During 2020, inspections further declined by more than 20% (OSHA). On a federal level, two-thirds fewer OSHA inspections were conducted during the first five months of the pandemic compared to the same period in 2019 (Fernández Campbell & Yerardi, 2020). In the current study, only 1 in 3 formal complaints (reported by a named employee) that required an inspection received one. This reduced inspection and enforcement activity may have limited the effectiveness of standards meant to help protect worker health and safety (Michaels & Wagner, 2020).

Alternatively, worker complaints may be a weak indicator of workplace risk of COVID-19 exposure, either because workers are not aware of the risks in their workplace or because they are hesitant or discouraged by employers to report workplace risks (Eidelson, 2020). Worker understanding of COVID-19 risk, awareness of the complaint process as a protected legal right, union membership, tenure with an employer, legal status, and fear of retaliation all contribute to propensity to file a complaint (Kalleberg, Wallace, & Althauser, 1981; Weil, 2007). For example, previous research has found a greater number of citations issued per employee among union compared to non-union companies (Scherer, Kaufman, & Ainina, 1993). Also inherent to any complaint system is the assumption that workers who experience a violation will complain (Weil & Pyles, 2005). Thus, higher rates of complaints may instead reflect the worker groups with greater empowerment, representation, resources, or capacity to advocate for better workplace protections. The possibility that Cal/OSHA complaints reflect empowerment rather than risky workplace environments highlights the need for *proactive* guideline dissemination, inspections, and enforcement beyond reactive protocols that respond to the workers that are most vocal rather than the most vulnerable.

While complaints were not indicative of the industries with the highest COVID-19 mortality rates, industries with the highest inspection rates did align with those with the highest mortality. For example, inspections were most common in Agriculture (21.5%) and comparatively less common in other industries (6.5-9.4%). Further research is needed to quantify COVID-19 mortality that can be causally linked to ill-addressed workplace citations.

Unlike inspections, no discernable pattern emerged between fines and COVID-19 mortality rates across industries. For instance, Health Care had the lowest mortality rate and received the highest total fines ($425,550) followed by Manufacturing ($304,850) which had the second highest mortality rate. This lack of correlation between OSHA fines and COVID-19 mortality risk aligns with prior literature showing that OSHA inspections with fines in manufacturing plants were linked to fewer injuries however the effectiveness declined from 1979-85 and became insignificant by 1992-98 (Gray & Mendeloff, 2005). Another study showed that inspections with fines specifically for personal protective equipment in manufacturing reduced injury from harmful exposures (Mendeloff & Gray, 2005), suggesting that employers may be more responsive to certain violations. These mixed results underscore our lack of understanding of if and how OSHA fines may influence workplace protections against COVID-19 exposure and mortality, meriting further examination of fines as an effective deterrent.

We also found that surges in worker complaint rates in spring and summer were followed by surges in COVID-19 mortality across industries, consistent with patterns previously documented by Hanage and colleagues (Hanage et al., 2020). This pattern did not prevail in the late fall surge however, which was not preceded by a notable increase in occupational complaints. The apparent temporal link between workplace complaints and mortality documented in the early months of the pandemic may simply reflect the overall temporal pattern of pandemic severity (CA Department of Public Health, 2021). Differences in the number of complaints filed across industries likely reflect the reopening of nonessential sectors. For example, summer complaints in Retail Trade were triple than those in the spring once nonessential businesses began to reopen, such as clothing stores and coffee shops.

Workplace exposure to COVID-19 is avoidable with adoption of protective equipment and other prevention measures, hence essential worker status does not imply an inevitable increase in COVID-19 mortality risk. Indeed, some essential worker groups in California experienced no increased mortality risk (Chen et al., 2021). In our data, industries with high COVID-19 death rates may indicate sectors or employers in need of information or technical assistance to support high-quality COVID-19 protection practices. Industries such as Agricultural and Manufacturing with low rates of complaints and high death rates may also identify groups needing better employee representation, education, or protection from retaliation. Similarly, we found that while complaints from workers in the health care sector accounted for the largest proportion and rate of all complaints, this sector also experienced one of the lowest death rates. Lower death rates among high-complaint industries suggests that workers empowered to take action to improve occupational safety can reduce mortality risk despite apparent high exposure.

Our descriptive study has notable considerations. Complaints filed to Cal/OSHA likely underrepresent the true number of COVID-19 employer violations. Similarly, deaths from COVID-19 are likely underestimated. However, the use of mortality rather than incidence avoids bias from underdiagnosis of COVID-19. In addition, lack of sufficient data on the number of California employees at each establishment precluded our ability to calculate establishment-specific rates of complaints, onsite inspections, and fines. Last, underlying conditions that worsen COVID-19 outcomes may differ between industries contributing to higher mortality (e.g., if diabetes diagnosis were more frequent among meatpacking workers than health care workers).

## Conclusions

In this novel study, we leveraged publicly available data of 2020 COVID-19-related Cal/OSHA complaints to evaluate whether complaints are an indicator of non-compliant industries and employers with workers at greater risk for COVID-19 exposure and mortality. Our analysis suggests that the Cal/OSHA complaint system warrants a closer evaluation to ensure that it is operating as intended: to “ensure safe and healthful working conditions for workers” (OSHA). Existing data suggest that even occupations with high COVID-19 exposure risk (e.g., health care workers) do not necessarily have elevated COVID-19 mortality risk. Rather, differences in COVID-19 infection and mortality risk likely exist in part because of inadequate adoption and enforcement of COVID-19-safe procedures in the workplace. This study suggests that complaints may be indicative of worker representation, power, and capacity to file a complaint within a given industry or establishment. If Cal/OSHA relies heavily on at-risk workers to complain about employer COVID-19 violations, future studies should examine which individual- and institutional factors determine who complains, what contributes to worker awareness of risk exposure, and what encourages worker participation in the complaint system for those with the greatest need of protection.

## Data Availability

All data are publicly available at the California Occupational Safety and Health Administration (Cal/OSHA) and the California Department of Public Health.

## Acknowledgments

We wish to thank Derek Wagner from StataCorp for assistance with data visualization.

## References

Brown, G. (2015). Malpractice by the Labor Movement: Relinquishing the Fight for Occupational Health and Safety in California. Paper presented at the New Labor Forum.

CA Department of Industrial Relations. Cal/OSHA safety and health complaint handling simplified process. Retrieved February 22, 2021. https://www.dir.ca.gov/dosh/caloshacomplaintflowchart.html.

CA Department of Industrial Relations. Citations for COVID-19 Related Violations. Retrieved February 22, 2021. https://www.dir.ca.gov/dosh/COVID19citations.html.

CA Department of Public Health. (2021). Tracking COVID-19 in California. Retrieved March 15, 2021. https://covid19.ca.gov/state-dashboard/.

Cal/OSHA. Recording and Reporting Requirements for COVID-19 Cases. Retrieved August 26, 2021. https://www.dir.ca.gov/dosh/coronavirus/Reporting-Requirements-COVID-19.html

Cal/OSHA. (2021a). COVID-19 Guidance and Resources. Retrieved April 17, 2021. https://www.dir.ca.gov/dosh/coronavirus/.

Cal/OSHA. (2021b). File a Workplace Safety Complaint. Retrieved March 31, 2021. https://www.dir.ca.gov/dosh/complaint.htm.

Cal/OSHA. (2021c). Interim Guidelines on Protectng Workers from COVID-19. Retrieved from https://www.dir.ca.gov/dosh/coronavirus/General-Industry.html..

Chen, Y.-H., Glymour, M., Riley, A., Balmes, J., Duchowny, K., Harrison, R., Bibbins-Domingo, K. (2021). Excess mortality associated with the COVID-19 pandemic among Californians 18-65 years of age, by occupational sector and occupation: March through October 2020. medRxiv.

Eidelson, J. (2020, August 27, 2020.). Covid Gag Rules at U.S. Companies Are Putting Everyone at Risk. Bloomberg Businessweek. Retrieved from https://www.bloomberg.com/news/features/2020-08-27/covid-pandemic-u-s-businesses-issue-gag-rules-to-stop-workers-from-talking.

Fernández Campbell, A., & Yerardi, J. (2020, August 18, 2020.). Fewer inspectors, more deaths: The Trump administration rolls back workplace safety inspections. Vox. The Center for Public Integrity. Retrieved from https://publicintegrity.org/politics/system-failure/deaths-cutbacks-workplace-safety-inspections-osha/.

Gray, W. B., & Mendeloff, J. M. (2005). The declining effects of OSHA inspections on manufacturing injuries, 1979–1998. ILR Review, 58(4), 571–587.

Hanage, W. P., Testa, C., Chen, J. T., Davis, L., Pechter, E., Seminario, P., Krieger, N. (2020). COVID-19: US federal accountability for entry, spread, and inequities—lessons for the future. European journal of epidemiology, 1–12.

Kalleberg, A. L., Wallace, M., & Althauser, R. P. (1981). Economic segmentation, worker power, and income inequality. American journal of sociology, 87(3), 651–683.

Mendeloff, J., & Gray, W. B. (2005). Inside the black box: How do OSHA inspections lead to reductions in workplace injuries? Law & Policy, 27(2), 219–237.

Mendeloff, J., & Seabury, S. A. (2013). Inspection Targeting Issues for the California Department of Industrial Relations Division of Occupational Safety and Health. Retrieved from

Michaels, D., & Wagner, G. R. (2020). Occupational Safety and Health Administration (OSHA) and worker safety during the COVID-19 pandemic. Jama.

NIOSH Industry and Occupation Computerized Coding System (NIOCCS). Retrieved March 14, 2021. https://csams.cdc.gov/nioccs/.

OSHA. About OSHA. Retrieved May 30, 2021. https://www.osha.gov/aboutosha#:∼:text=OSHA’s%20Mission,%2C%20outreach%2C%20education%20and%20assistance.

OSHA. Inspections within Industry. Retrieved March 31, 2021. https://www.osha.gov/pls/imis/industry.html.

Scherer, R. F., Kaufman, D. J., & Ainina, M. F. (1993). Resolution of complaints by OSHA in union and non-union manufacturing organizations. Journal of Applied Business Research (JABR), 9(2), 55–61.

State of California. (2020a). Essential Workforce. Retrieved March 26, 2021. https://covid19.ca.gov/essential-workforce/

State of California. (2020b). Executive Order N-33-20: State of Emergency to exist in California as a result of the threat of COVID-19.. Retrieved March 20, 2021. https://covid19.ca.gov/stay-home-except-for-essential-needs/.

The New York Times. (2021). Coronavirus in the U.S.: Latest Map and Case Count. Retrieved March 31, 2021. https://www.nytimes.com/interactive/2020/us/coronavirus-us-cases.html#states.

US Census Bureau. North American Industry Classification System.. Retrieved January 31, 2021. https://www.census.gov/naics/?48967.

Waltenburg, M. A., Rose, C. E., Victoroff, T., Butterfield, M., Dillaha, J. A., Heinzerling, A., Fedak, K. M. (2021). Coronavirus disease among workers in food processing, food manufacturing, and agriculture workplaces. Emerging Infectious Diseases, 27(1), 243.

Weil, D. (2007). Exploring the complaints and compliance gap under US workplace policies. LERA For Libraries.

Weil, D., & Pyles, A. (2005). Why complain-complaints, compliance, and the problem of enforcement in the us workplace. Comp. Lab. L. & Pol’y. J., 27, 59.

