## Appendices for "A descriptive analysis of 2020 California Occupational Safety and Health Administration COVID-19-related complaints"

**Appendix A**. Industry and Complaint Variable Coding

**Industry**: variable coding using NAICS industry name
1=Health Care and Social Assistance
2=Retail Trade
3=Manufacturing
4=Accommodation and Food Services
5=Transportation and Warehousing
6=Agriculture, Forestry, Fishing and Hunting
7=Other (Public Administration; Construction; Wholesale Trade; Other Services (except Public Administration); Management of Companies and Enterprises; Professional, Scientific, and Technical Services; Educational Services; Arts, Entertainment, and Recreation; Finance and Insurance; Real Estate and Rental and Leasing; Information; Utilities; Mining, Quarrying, and Oil and Gas Extraction)

**Business Type**: variable coded using Cal/OSHA "ownership_type"
1 = local government
2 = private sector
3 = state government

**Complaint Type**: variable coded using Cal/OSHA "severity_subject"
1 (Health) = Discrimination O-Health, I-Health, O-Health, Discrimination S-Health
2 (Safety) = O-Safety, I-Safety, S-Safety
3 = (Health & Safety) = O-Health;O-Safety, O-Health;S-Health, O-Safety;S-Health, S-Health;S-Safety

**Complaint Severity**: variable coded using Cal/OSHA "severity_subject"
1 (Low) = Discrimination O-Health, I-Health, I-Safety, O-Health, O-Safety, O-Health;O-Safety
2 (High) = O-Health;S-Safety, O-Safety;S-Health, Discrimination S-Health; S-Health;S-Safety

**Formality**: variable coded using Cal/OSHA "formality"
1 = formal
2 = nonformal

**Inspection**: variable coded using Cal/OSHA "do_insp"
0 (No) = do_insp = N; NA
2 (Yes) = do_insp = Y

**Reason for Inspection**: variable coded using Cal/OSHA "reason_for_insp" and restricted to "do_insp=Y"
1 (Required) = Valid Formal Complaint Submitted; Formal Complaint Alleges Recordkeeping Deficiencies; Alleged Imminent Danger
2 (Follow-up) = Inadequate/No ER Response to Injury - Includes Employee Dispute; Inspection of Employer is Scheduled/in Progress; Abatement Follow-up/Monitoring Inspection
3 (AD Discretion) = AD Discretion

**Penalty**: variable coded using Cal/OSHA "proposed_penalty" and restricted to "do_insp=Y"
0 (No) = proposed_penalty=0
1 (Yes) = proposed_penalty>0

**Appendix B**. Distribution of Cal/OSHA COVID-19-related inspections characteristics by industry, 2020.

| | Total | Health Care & Social Assistance | Retail Trade | Manufacturing | Accommodation & Food Services | Transportation & Warehousing | Agriculture, Forestry, Fishing & Hunting | Other | P-value |
|---|---|---|---|---|---|---|---|---|---|
| | *row n (%)* | | | | | | | | |
| Inspections | | | | | | | | | 0.000 |
| Total | 627 (100.0) | 153 (24.4) | 98 (15.6) | 81 (13.0) | 63 (10.0) | 41 (6.5) | 32 (5.1) | 159 (25.4) | |
| | | | | | *column n (%)* | | | | |
| Business Type | | | | | | | | | 0.000 |
| Local | 22 (3.5) | 4 (2.6) | 0 (0.0) | 0 (0.0) | 0 (0.0) | 3 (7.3) | 0 (0.0) | 15 (9.4) | |
| Private | 587 (93.6) | 146 (95.4) | 98 (100.0) | 81 (100.0) | 63 (100.0) | 37 (90.3) | 32 (100.0) | 130 (81.8) | |
| State | 18 (2.9) | 3 (2.0) | 0 (0.0) | 0 (0.0) | 0 (0.0) | 1 (2.4) | 0 (0.0) | 14 (8.8) | |
| Complaint Type | | | | | | | | | 0.062 |
| Health (H) | 560 (89.3) | 141 (92.2) | 89 (90.8) | 67 (82.7) | 61 (96.8) | 37 (90.2) | 30 (93.8) | 135 (84.9) | |
| Safety (S) | 28 (4.5) | 2 (1.3) | 4 (4.1) | 5 (6.2) | 1 (1.6) | 2 (4.9) | 0 (0.0) | 14 (8.8) | |
| H & S | 39 (6.2) | 10 (6.5) | 5 (5.1) | 9 (11.1) | 1 (1.6) | 2 (4.9) | 2 (6.2) | 10 (6.3) | |
| Reason | | | | | | | | | 0.000 |
| Required | 266 (42.4) | 66 (43.1) | 36 (36.7) | 29 (35.8) | 17 (27.0) | 23 (56.1) | 13 (40.6) | 82 (51.6) | |
| Follow-up | 101 (16.1) | 17 (11.1) | 24 (24.5) | 8 (9.9) | 20 (31.7) | 6 (14.6) | 0 (0.0) | 26 (16.3) | |
| AD's Discretion | 260 (41.5) | 70 (45.8) | 38 (38.8) | 44 (54.3) | 26 (41.3) | 12 (29.3) | 19 (59.4) | 51 (32.1) | |
| Formality | | | | | | | | | 0.376 |
| Formal | 245 (39.1) | 59 (38.6) | 32 (32.7) | 32 (39.5) | 20 (31.7) | 20 (48.8) | 12 (37.5) | 70(44.0) | |
| Nonformal | 382(60.9) | 94 (61.4) | 66 (67.3) | 49 (60.5) | 43 (68.3) | 21 (51.2) | 20 (62.5) | 89 (56.0) | |
| Severity | | | | | | | | | 0.000 |
| Low | 500 (79.7) | 91 (59.5) | 88 (89.8) | 72 (88.9) | 56 (88.9) | 37 (90.2) | 27 (84.4) | 129 (81.1) | |
| High | 127 (20.3) | 62 (40.5) | 10 (10.2) | 9 (11.1) | 7 (11.1) | 4 (9.8) | 5 (15.6) | 30 (18.9) | |
| Penalty | | | | | | | | | 0.005 |
| No | 595 (94.9) | 138 (90.2) | 95 (96.9) | 75 (92.6) | 63 (100.0) | 38 (92.7) | 29 (90.6) | 157 (98.7) | |
| Yes | 32 (5.1) | 15 (9.8) | 3 (3.1) | 6 (7.4) | 0 (0.0) | 3 (7.3) | 3 (9.4) | 2 (1.3) | |

*Note: Abbreviation: AD = Area Director; Chi-square test compares complaint characteristic across the six industry categories.*
